# Efficacy and Safety of PD-1/PD-L1 plus CTLA-4 antibodies +/− Other Therapies in Lung Cancer: A Systematic Review and Meta-Analysis

**DOI:** 10.1101/2020.01.08.20016915

**Authors:** Xiang Shen, Hua Xiao, Shangke Huang, Jiexing Liu, Zhuolan Ran, Bin Xiong

## Abstract

**Background:** Immune checkpoint inhibitors have brought hope for patients with advanced lung cancer, among which PD-1/PD-L1 and CTLA-4 inhibitors have achieved considerable results. This meta-analysis aims to investigate the efficacy and safety of PD-1/PD-L1 combined with CTLA-4 antibodies in patients with advanced lung cancer.

**Materials and Methods:** Randomized controlled trials (RCTs) study of PD-1/PD-L1 combined with CTLA-4 inhibitors for advanced lung cancer was searched in the database for systematic review and meta-analysis.

**Results:** The meta-analysis finally included 4 trials (actually 3 trials), and the results showed that the overall response rate of PD-1/PD-L1 combined with CTLA-4 inhibitor was higher than that of the control group (ORR = 2.29 [95% CI 1.58 3.32], P <0.0001). PFS significantly improved compared with conventional treatment (HR = 0.69 [95% CI 0.56, 0.85], P = 0.0005), which was statistically significant. OS also improved but did not statistically significant (HR = 0.80 [95% CI 0.61, 1.05], P = 0.11). PD-1/PD-L1 combined with CTLA-4 inhibitors had a higher incidence of adverse reactions than conventional treatment (OR = 1.72 [95% CI 0.59, 5.09], P = 0.33), but the incidence of grade≥3 AEs was lower than the control group (OR = 0.96 [95% CI 0.64, 1.44], P = 0.85).

**Conclusion:** Double checkpoint inhibitor PD-1/PD-L1 combined with CTLA-4 antibody has synergistic anti-cancer activity and improved ORR and PFS in lung cancer. Adverse reactions are more common than conventional treatment, but are generally controllable. Therefore, PD-1/PD-L1 combined with CTLA-4 inhibitors provides a beacon for the treatment of advanced lung cancer, which has guiding significance for the treatment selection in the future.

## Introduction

Although undergoing smoking cessation, early screening and treatment have reduced lung cancer mortality,^1^ it still remains the most deadly cancer among cancer patients. Because lung cancer in the early stages is usually asymptomatic or has only cough and sputum without attention, most patients are diagnosed in the middle-late stages and haven’t opportunity for surgery. It analysised the 5-year survival rate of patients with stage IV is only 4% in the Cancer (2019).^2^ Even with the presence of treatable gene mutations, using targeted drugs will eventually activate acquired resistance mutations or compensatory pathways,^3^ leading to disease progression occurring 8-19 months after starting treatment.^4 5^ We need to actively find other therapies to solve this dilemma.

Studies have found that cancer cells in the context of the tumor microenvironment (TME) through regulatory T cells (Tregs) change immune homeostasis, suppress the activation and effector functions of innate and acquired immune systems, thereby escape immune surveillance.^6 7^ When PD-1 is combined with PD-L1, it inhibits the proliferation and survival of CD8 + toxic T cells;^8^ at the same time, changes T cell differentiation, impairs the differentiation of effector T cells (Teff) and memory T cells (Tm), up-regulates regulatory T lymphocytes (Treg) and depleted T cells (Tex), thus suppress T-cell immune effects. CTLA-4 has greater affinity than CD28 for binding to CD80 and CD86 on antigen-presenting cells, reducing the synergistic stimulation of the T cell receptor signal,^9^ thereby inhibiting CD28-B7-1/2 costimulation, leading to T cell dysfunction and apoptosis.^10^ This is the escape of adaptive immunity to endogenous anti-tumor mechanisms.^11^ Immune checkpoint inhibitors (ICIs) restore and maintain the function of the immune system against tumor cells by blocking specific signaling pathways, in which cytotoxic T cells play a important role in immune monitoring and anti-tumor responses.^12^ Compared with chemotherapy, PD-1/PD-L1 and CTLA-4 inhibitors have higher objective response rate or longer overall survival and better toxicity profile.^13 14^ In particular, patients without targeted cancer mutation genes receiving tumor immunotherapy is improved the long-term survival rate,^15 16^ and it is a new treatment standard for patients with advanced non-small cell lung cancer with new diagnosis, general conditions, and no contraindications.^17^

However, mono-immunotherapy significantly prolonged survival in patients with high PD-L1,^18^ which is only beneficial to a small number of patients.^19^ Greater therapeutic effectiveness can be achieved at the cost of corresponding side effects through synergistic or combined treatment. Recent clinical trials have demonstrated that the dual checkpoint inhibitors Ipilimumab plus Nivolumab can synergistically enhance T cell functions and the immune system’s ability to fight tumor cells through different and complementary mechanisms,^20^ have a better curative effect than Nivolumab monotherapy. The effect was better in patients with high PD-L1,^21^ and reducing the proportion of patients with advanced tumors.^22^

Immune-related adverse events (irAEs), which are different from chemotherapy-related toxic reactions,^23^ are up-regulated immune systems leading to severe inflammatory reactions and immune toxicity.^24^ The mechanism may be related to Tregs. Tregs are one of the most abundant suppressor cells in the TME, and express a large number of checkpoint molecules such as CTLA-4 and PD-1, to maintain immune homeostasis and avoiding autoimmunity. Therefore, the targeting effect of ICIs on Tregs may lead to the occurrence of irAEs.^25^ These events are usually controllable^26^ and can be effectively controlled and managed by immunosuppressive methods such as steroids.^27^ However, there are still a few patients happen serious adverse reactions, with the highest mortality caused by neurological and cardiac toxicity.^28^

Several clinical trials have been conducted to evaluate the efficacy and toxicity of PD-1/PD-L1 combined with CTLA-4 antibodies in patients with advanced lung cancer, but the results such as progression-free survival (PFS) and overall survival(OS) are controversial. Therefore, we conducted a systematic meta-analysis using data from published literature to make a systematic review and meta-analysis on the efficacy and safety of PD-1/PD-L1 combined with CTLA-4 antibody and/or other therapeutic regiments in lung cancer.

## Materials and methods

We follow principle in this systematic review and meta-analysis study is the system review and meta-analysis (PRISMA) guidelines for priority reporting projects.

### Systematic Review and Meta-Analysis of All RCTs Testing the PD-1/PD-L1 plus CTLA-4 antibodies +/− Other Therapies in lung cancer

#### Data sources and search strategy

We searched PubMed, Web of Science, the Cochrane Library, Ovid, Embase for RCTs testing the PD-1/PD-L1 plus CTLA-4 antibodies in patients with lung cancer, without any language or date restrictions. The following retrieval strategy was employed: ((((PD-1 OR PD-L1 OR Pembrolizumab OR Nivolumab OR Cemiplimab-rwlc OR Toripalimab OR Sintilimab OR Atezolizumab OR Avelumab OR Durvalumab) AND (CTLA-4 OR Ipilimumab OR Tremelimumab)) AND lung cancer) AND trial). And manual searches were performed for references that met the inclusion criteria to avoid missing any studies.

#### Inclusion and exclusion criteria

Inclusion and exclusion criteria were independently evaluated by two researchers based on the PICO principle. The included studies must meet the following criteria: (1) studies published in English; (2) randomized controlled clinical trials to study the efficacy and safety of (PD-1 OR PD-L1 OR Pembrolizumab OR Nivolumab OR Cemiplimab-rwlc OR Toripalimab OR Sintilimab OR Atezolizumab OR Avelumab OR Durvalumab) AND (CTLA-4 OR Ipilimumab OR Tremelimumab); (3) the patients were histologically diagnosed with lung cancer,not including HIV, organ transplantation, viral infection and other special patients; (4) analysis related data of objective response rate(ORR), progression free survival (PFS), overall survival (OS), and toxicity. The following studies were excluded: (1) reviews, editorials, case reports, or animal and cellular studies; (2) incomplete required data; (3) no dose and usage.

#### Data extraction and quality assessment

Two researchers independently conducted the study selection process based on the inclusion and exclusion criteria, and extracted the following information: first author name, publication year, trial phase, clinical trial number, trial design, pathological type, sample size, usage and dosage, follow-up time, median OS and PFS, irAEs. If there are disagreements in the studies selection and data extraction process, reach a consensus through discussion or when necessary consult the third investigator.

We used the reporting method of Jadad et al^29^ to evaluate the quality of the included studies.

#### Data synthesis and statistical analysis

We used Review Manager 5.3 for statistical analysis. The Q test and I^2^ statistics were evaluated the statistical heterogeneity. When P<0.05 or I^2^>50%, heterogeneity is indicated statistically significant, and performed using random effects models, otherwise performed using fixed effect model. HRs>1 of OS and PFS was beneficial to the control group, while HRs<1 was beneficial to the PD-1/PD-L1 combined CTLA-4 antibody group. ORs>1 of ORR and AEs means higher effective rate and toxicity, while ORs<1 have lower effective rate and safety. P<0.05 was considered statistically significant, and all P values were bilateral.

## Results

### Study characteristics and risk of bias

Figure 1 shows a flow chart containing the study. A preliminary literature search screened a total of 3983 records from the database; 3317 of them were after deleting duplicate records; 86 were after screening; 43 were after screening titles and abstracts; 3 studies After reviewing each publication. All relevant references have been reviewed. Finally, three randomized controlled trials were included in this meta-analysis (one of the trials had two trials to compare the efficacy and toxicity with the control group, which were conducted as two independent trials), all using the response evaluation standard (RECIST) or WHO standards. All trials are randomized, controlled, open-label clinical trials. These included 430 experimental groups and 375control groups,^30-32^ which mainly studied Ipilimumab + Nivolumab and Durvalumab + Tremelimumab for the treatment of lung cancer. Table I summarizes the detailed characteristics of the included studies.

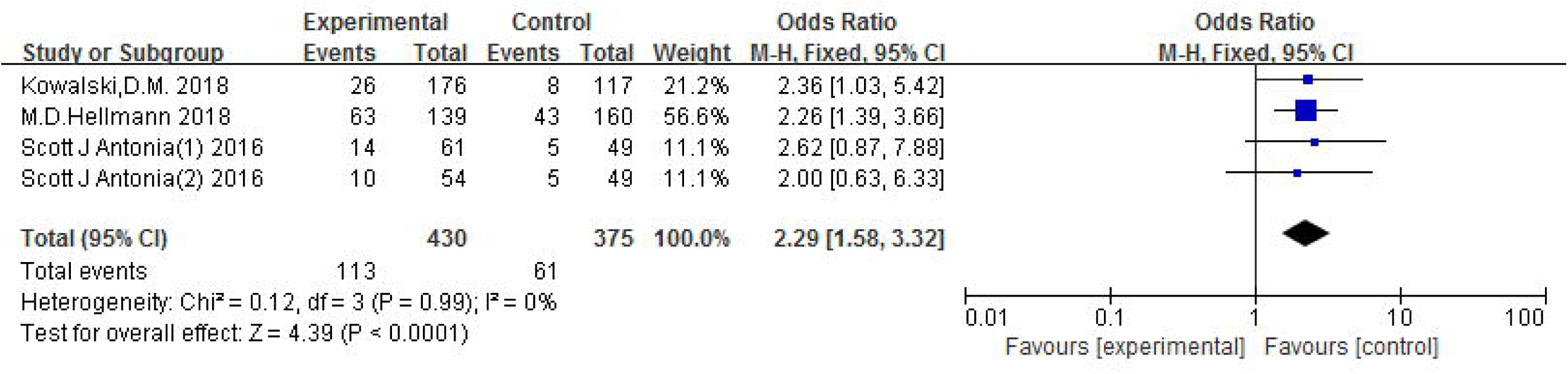

**Table I.**
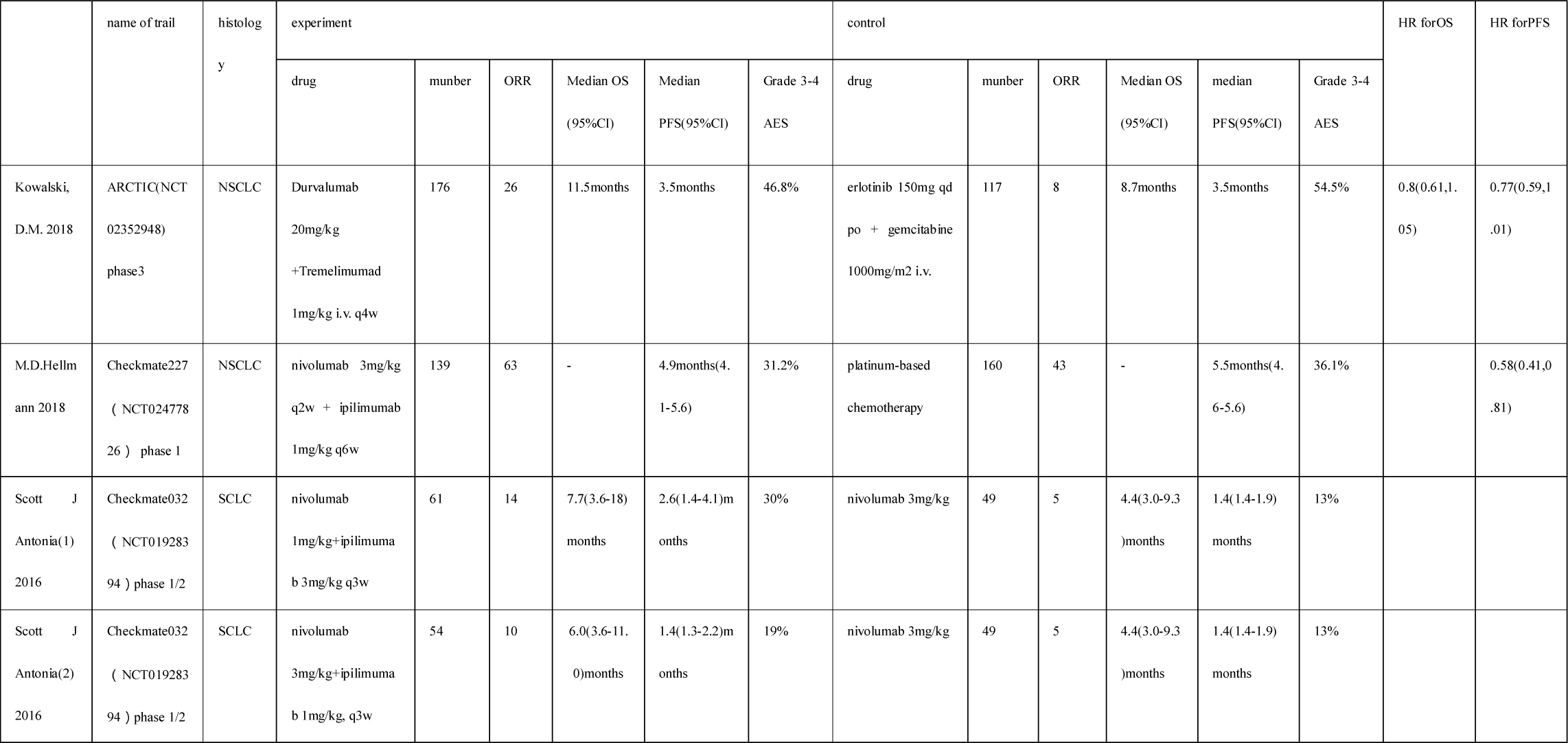

|  | name of trail | histology | experiment |  |  |  |  |  | control |  |  |  |  |  | HR for OS | HR for PFS |
| --- | --- | --- | --- | --- | --- | --- | --- | --- | --- | --- | --- | --- | --- | --- | --- | --- |
|  |  |  | drug | number | ORR | Median OS<br>(95% CI) | Median<br>PFS(95% CI) | Grade 3-4<br>AES | drug | number | ORR | Median OS<br>(95% CI) | Median<br>PFS(95% CI) | Grade 3-4<br>AES |  |  |
| Kowalski,<br>D.M. 2018 | ARCTIC(NCT02352948)<br>phase 3 | NSCLC | Durvalumab<br>20mg/kg<br>+Tremelimumab<br>1mg/kg i.v. q4w | 176 | 26 | 11.5months | 3.5months | 46.8% | erlotinib 150mg qd<br>po + gemcitabine<br>1000mg/m2 i.v. | 117 | 8 | 8.7months | 3.5months | 54.5% | 0.8(0.61,1.05) | 0.77(0.59,1.01) |
| M.D.Hellmann 2018 | Checkmate227<br>( NCT02477826 ) phase 1 | NSCLC | nivolumab 3mg/kg<br>q2w + ipilimumab<br>1mg/kg q6w | 139 | 63 | - | 4.9months(4.1-5.6) | 31.2% | platinum-based<br>chemotherapy | 160 | 43 | - | 5.5months(4.6-5.6) | 36.1% |  | 0.58(0.41,0.81) |
| Scott J<br>Antonia(1)<br>2016 | Checkmate032<br>( NCT01928394 ) phase 1/2 | SCLC | nivolumab<br>1mg/kg+ipilimumab<br>3mg/kg q3w | 61 | 14 | 7.7(3.6-18)months | 2.6(1.4-4.1)months | 30% | nivolumab 3mg/kg | 49 | 5 | 4.4(3.0-9.3)months | 1.4(1.4-1.9)months | 13% |  |  |
| Scott J<br>Antonia(2)<br>2016 | Checkmate032<br>( NCT01928394 ) phase 1/2 | SCLC | nivolumab<br>3mg/kg+ipilimumab<br>1mg/kg, q3w | 54 | 10 | 6.0(3.6-11.0)months | 1.4(1.3-2.2)months | 19% | nivolumab 3mg/kg | 49 | 5 | 4.4(3.0-9.3)months | 1.4(1.4-1.9)months | 13% |  |  |

We assessed the quality of each study in this analysis based on the Jadad score, and Table II provides the risk results of the bias assessment.

**Table II.** Risk of Bias

|  | random sequence generation (selection bias) | allocation concealment (selection bias) | blinding of participants and personnel (performance bias) | blinding of outcome assessment (detection bias) | incomplete outcome data (attrition bias) | selective reporting (reporting bias) | other bias |
| --- | --- | --- | --- | --- | --- | --- | --- |
| Kowalski D.M. 2018 | - | ? | - | ? | - | ? | + |
| M.D.Hellmann 2018 | - | ? | - | ? | - | ? | + |
| Scott J Antonia(1) 2016 | - | ? | - | ? | + | ? | + |
| Scott J Antonia(2) 2016 | - | ? | - | ? | + | ? | + |

### Overall response rate (ORR)

Finally, 4 studies conducted in 430 experimental arm patients and 375 control arm patients met the inclusion criteria and were included in the ORR analysis. The ORR funnel plot did not show significant asymmetry (Figure 3), and heterogeneity between studies was not significant (P= 0.99, I^2^=0%). The pooled OR for ORR was performed fixed effect model. This meta-analysis showed that ORR was significantly improved in the treatment of PD-1/PD-L1 combined with CTLA-4 antibody (OR=2.29[95%CI 1.58, 3.32], P<0.0001) (Figure 2), which was statistically significant. The clinical efficacy of PD-1/PD-L1 combined with CTLA-4 antibodies in patients with advanced lung cancer is higher than that in the control group. Regardless of whether it is NSCLC or SCLC, the results show that dual ICI therapies improves ORR.

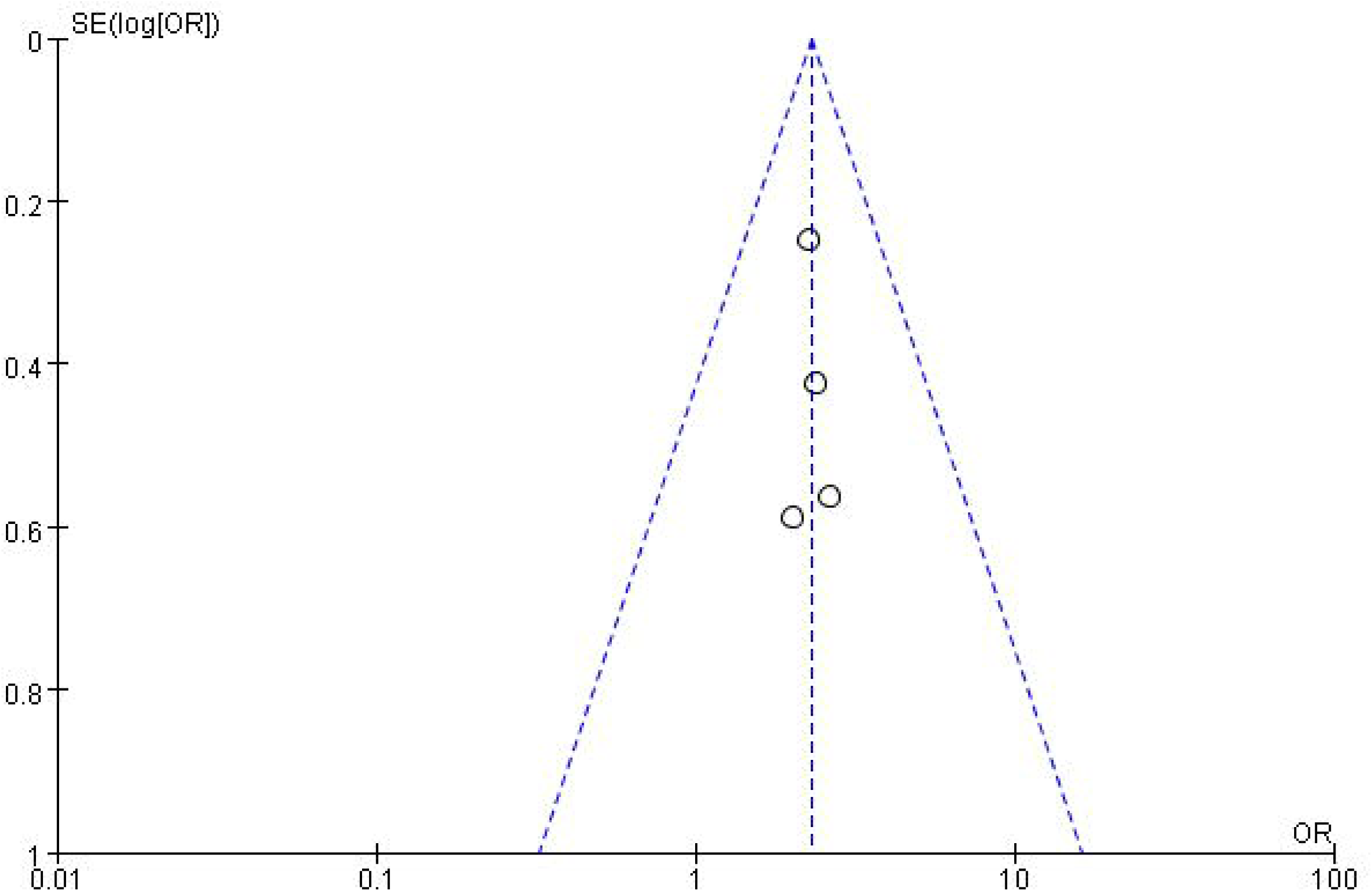

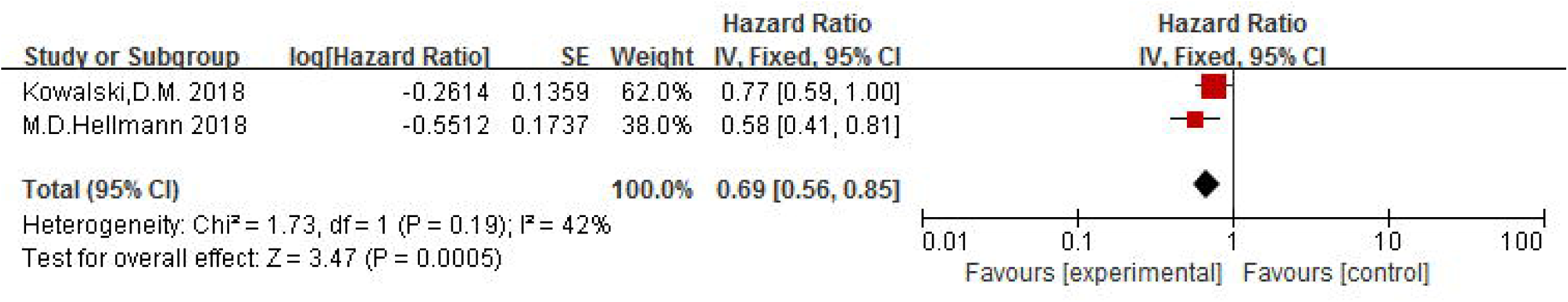

### progression-free survival (PFS)

Two trials were included for PFS evaluation. The funnel plot of PFS did not show significant asymmetry, and heterogeneity among studies was not significant (P = 0.19, I^2^ = 42%). The pooled HR for PFS was performed fixed effect model. In the treatment of lung cancer with PD-1/PD-L1 combined with CTLA-4 antibodies, the HR of PFS improved statistically (HR = 0.69 [95% CI 0.56, 0.85], P = 0.0005) (Figure 4).

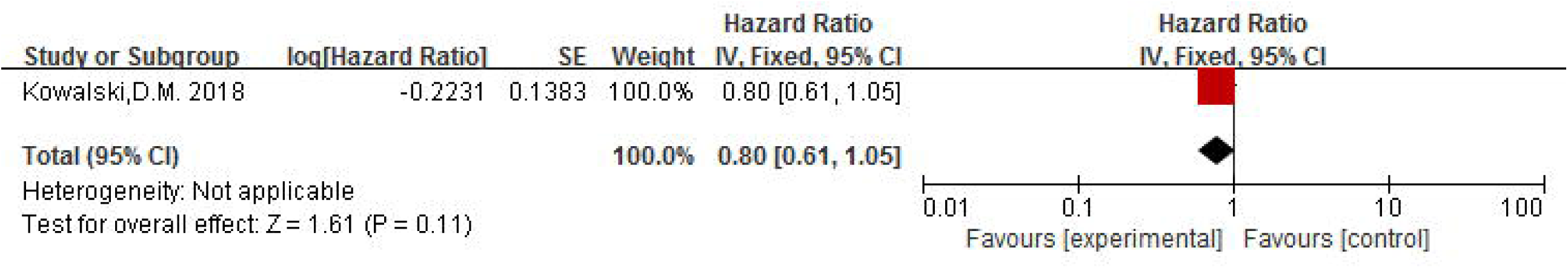

### overall survival (OS)

Only one trial reported OS combined HRs and 95% CI, and heterogeneity could not be evaluated. The results showed that the OS of PD-1/ PD-L1 combined with CTLA-4 antibody was improved but not statistically significant (HR = 0.80[95% CI 0.61, 1.05], P = 0.11) (Figure 5).

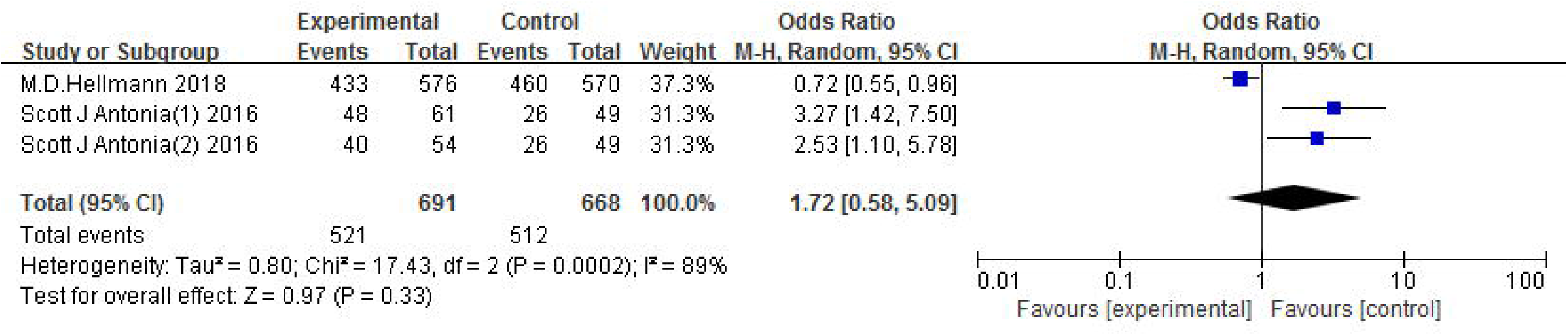

### Immune-related adverse events (irAEs)

Data from AEs were found in 3 studies with significant heterogeneity (P = 0.0002, I^2^ = 89%). The pooled OR for AEs was performed random effect model. The incidence of AEs was higher than the control group (OR = 1.72 [95% CI 0.59, 5.09], P = 0.33) (Figure 6), and fatigue was the most common. There are 4 studies with data from severe AEs (Grade ≥ 3) and heterogeneity (P = 0.1, I2 = 52%). The pooled OR for AEs (Grade≥3) was performed random effect model. The incidence of AEs (Grade ≥ 3) was lower than the control group (OR = 0.96[95%CI 0.64, 1.44], P = 0.85) (Figure 7), but it was not obvious. Among them, rash and diarrhea were the most common, anemia and neutrophilic granulocyte counts were most common during chemotherapy.

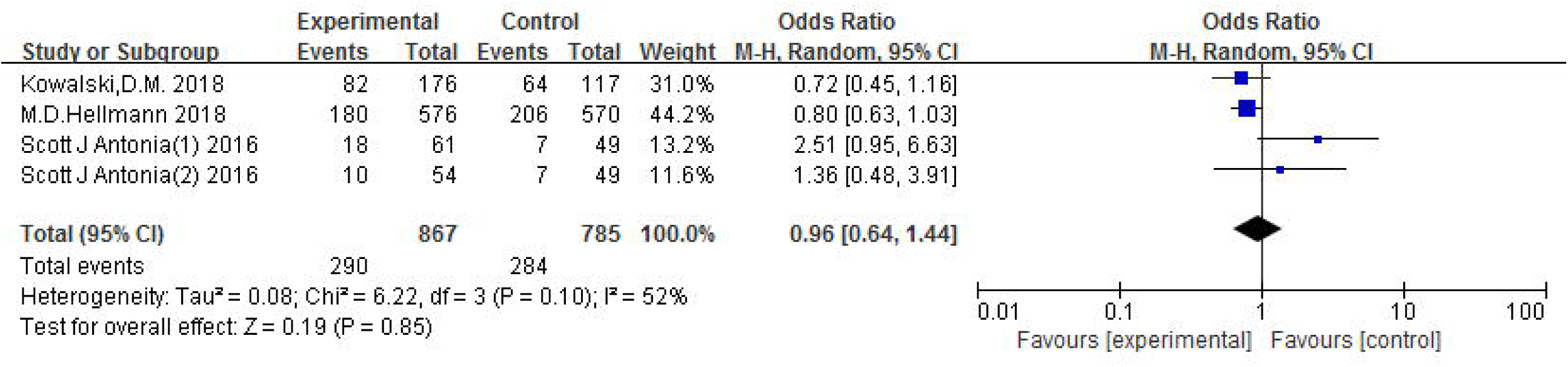

## Discussion

Most lung caner patients receive chemotherapy and/or radiation therapy,^2^ which have poor antitumor effects. In recent years, some clinical trials have achieved satisfactory results in the treatment of lung cancer with PD-1/PD-L1 and CTLA-4 antibodies, of which Pembrolizumab and Nivolumab can prolong survival for patients with advanced NSCLC,^33 34^ Durvalumab combined with chemotherapy can effectively improve the prognosis of patients with ES-SCLC.^35^ This meta-analysis investigates the efficacy of PD-1/PD-L1 plus CTLA-4 inhibitors for lung patients.

Nivolumab and Pembrolizumab mainly act on activated T cells in peripheral blood or tumors. By blocking the binding of PD-1 and PD-L1, they cancell the apoptosis of T cells and enable their normal activation and proliferation. Ipilimumab mainly acts on lymph nodes, by blocking the binding of the domain of CTLA-4 to its ligand, activate, proliferate and infiltrate T lymphocytes, and increase the level of active T cells in peripheral blood.^36^ However, less than 10% of CD8+ T cells in tumors have the ability to recognize cancer cells,^37^ and the anticancer effect may be caused by T cells entering the tumor from peripheral blood after treatment.^38^ Therefore, it is particularly important to increase the number of T cells and enhance the effector function through different mechanisms by combining PD-1/PD-L1 and CTLA-4 inhibitors. The combined blocking of CTLA-4 and PD-1/PD-L1 pathways increases the ratio of Teff to Tregs and myeloid-derived suppressor cells (MDSC, which significantly suppresses the ability of cells to respond), thereby reducing immunosuppression and promoting inflammation in the tumor microenvironment.^39^ At the same time, anti-CTLA-4 antibodies have antibody-dependent cell-mediated cytotoxicity, reduce the negative effect of PD-1 antibodies on activating Tregs; CTLA-4 antibodies lead to compensatory overexpression of tumor PD-L1, and PD-1 antibodies block the binding of PD-L1 to PD-1; CTLA-4 antibodies increase activated T cells and Tm in peripheral blood, and PD-1 antibodies relieve the inhibition of T cell anti-cancer activity by tumors. Our results reveal the significant effect of dual checkpoint inhibitors treat for advanced lung cancer, with significant improvements in ORR (OR=2.29[95%CI 1.58, 3.32], P<0.0001) and PFS (HR = 0.69 [95% CI 0.56, 0.85], P = 0.0005); OS also improves (HR = 0.80[95% CI 0.61, 1.05], P = 0.11), but no statistical significance.

Can we combine immunosuppressants with other treatments for lung cancer? By activating aKT, cisplatin rapidly increases the expression of PD-L1 in the tumor microenvironment in a dose-dependent manner, increases the clinical efficacy of immunotherapy,^40 41^ and does not affect the immune background of NSCLC after induction, and does not significantly damage antitumor Immune,^42^ but may not persistently. Radiotherapy can restore the recruitment of Th1 lymphocytes in the tumor microenvironment,^43^ stimulate antigen presentation, increase tumor antigenicity,^44^ form an “immune center”, promote tumor immune response.^45 46^ Through more effective control of the immune system, local therapy is converted into systemic therapy, which makes it have synergistic and more effective activity on tumor cells.^47^ Regarding when to use combination therapy, a meta-analysis by Zhou et.al suggests patients with large tumor volume should be treated with immunity combined with other therapies to produce deeper and long-term efficacy, while patients with low tumor volume or extremely high PD-L1 TPS should be treated with immunomonotherapy.^48^ which requires more clinical trials validation.

In order to expand the population benefiting from immunotherapy, detection of biomarkers before treatment is necessary. In the Checkmate 568 study, patients with stage IIIB/IV NSCLC received Nevolumab and Ipilimumab. The ORs of patients with PD-L1 expression < 1% and ≥ 1% were 14.9%[95% CI 8.9, 22.8], 41.3%[95% CI 33,50], respectively, MPFS is 2.8months[95% CI 2.1,4.0], 6.8months[95% CI 4.0, 11.3], and the OR of patients with PD-L1 ≥ 50% is 50%[95% CI 37.6, 62.4].^49^ It is suggested that tumor PD-L1 expression is significantly enhanced when receiving dual immunosuppressants at 1% or higher, which has been considered to a predictive biomarker of immune checkpoint inhibitor response.^50^ However, PD-L1 expression is heterogeneous and is affected by chemotherapy and targeted therapy.^51^ The immune response to anti-CTLA-4 or anti-PD-1/CTLA-4 combination therapy in high TMB tumors may not depend on PD-L1 expression.^52^ Several studies have shown a correlation between TMB and the benefits of ICIs in NSCLC, SCLC, and melanoma.^53^ The higher the TMB, the more mutations in the tumor genome, the higher the probability of new antigens appearing on the tumor surface, the stronger the immunogenicity, making the tumor more easily recognized by T cells;^54^ and it is significantly related to the response of immunotherapy.^55^ Hellmann et al. according to TMB quantitative classification: low (0 to <143 mutations), medium (143 to 247 mutations) and high (≥248 mutations), the ORR of Nivolumab+ipilimumab in SCLC is 22.2%, 16.0%, 46.2%, MPFS is 1.5 months[95% CI 1.3, 2.7], 1.3 months[95% CI 1.2, 2.1], 7.8 months[95%CI 1.8, 10.7], MOS is 3.4 months[95% CI 2.8,7.3], 3.6 months[95% CI 1.8,7.7], 22.0 months[95% CI 8.2, NR], suggesting that dual immunosuppressants have better therapeutic effect and larger clinical value in patients with high tumor mutation load.^56^ The expression of PD-L1 and TMB can be used as predictors of immunotherapy.

The related adverse reactions of PD-1/PD-L1 combined with CTLA-4 inhibitors cannot be ignored. The risk of irAEs in patients treated with CTLA-4 is dose-related,^57^ most grade 3 or higher irAES occurs within 8-12 weeks after starting treatment; PD-1 antibody-related irAEs are relatively low in frequency, most irAEs occur within the first 6 months of treatment and take longer than treatment of CTLA-4 related toxicity.^58^ The organ involvement spectrum of the two is also different, anti-PD-1 drugs cause arthritis more frequently, and anti-CTLA-4 drugs are more related to colitis.^59^ Combining anti-CTLA-4 antibodies and anti-PD-1 antibodies can increase the incidence and severity of irAEs and occur earlier.^60^ This is consistent with our research finding. A big data analysis showed that the risk of immune events is associated with TMB. Cancers with high TMB, such as NSCLC, are associated with higher irAEs during anti-PD-1 therapy.^61^ Receiving low-dose glucocorticoids does not inhibit the immune response caused by ICIS, and can continue and maintain a beneficial response to ICIs.^59^ The combination of anti-PD-1 antibodies and immune antibodies of selectively target specific inflammatory mediators can prevent or delay the progression of advanced tumors with autoimmune diseases without affecting the anti-tumor effect of anti-PD-1 antibodies.^62^ It was observed that even if patients receiving PD-1 antibodies stopped treatment due to the reaction, they could have a longer duration of action without the need to rush to the next treatment.^63^ However, long-term exposure to immunosuppressants can lead to rare fatal immune-related events.^64 65^

In our meta-analysis, we included 4 randomized controlled clinical trials, the results showed that the combination of PD-1/PD-L1 and CTLA-4 antibody was effective for the treatment of lung cancer and the occurrence of adverse reactions is controllable.It provides a good option for the treatment of SCLC. A non-randomized controlled trial of Ipilimumab and Nivolumab given postoperatively to patients with ES-SCLC was better than the control group compared with OS-1 year.^66^ Contrary to our findings, more randomized, multicenter, and large sample studies are needed to confirm and evaluate the cost of treatment strategies and patients’ quality of life. Our research also has certain limitations that may affect the final results. First, most studies have a small sample size, and currently there are very few data on dual-immune checkpoint inhibitor combination therapy, and fewer studies involving randomized or blinded methods. Second, we extracted trial data from published articles but did not have the patient’s original data, which could lead to biases in data analysis. Therefore, more large-scale clinical trials are needed to further verify the effectiveness and safety of dual immunosuppressive antitumor therapy. Multiple RCTs are currently underway to compare the efficacy of PD-1/PD-L1 in combination with CTLA-4 inhibitors and/or other treatment options for advanced cancer, Durvalumab + Tremelimumab (MYSTIC/NCT02453282,^67^ ADRIATIC/NCT03703297,^68^ NEPTUNE/NCT02542293^69^), Durvalumab + Tremelimumab + Chemotherapy (POSEIDON/NCT03164616,^70^ CASPIAN/NCT03043872^71^), Nivolumab + Ipilimumab (CheckMate227/NCT02477826,^72^ CheckMate816/NCT02998528^73^), Pembrolizumab + Ipilimumab (KEYNOTE-598/NCT03302234^74^). It is necessary to pay close attention to the test results, perform update analysis, and conduct long-term follow-up.

In conclusion, PD-1/PD-L1 inhibitors combined with CTLA-4 inhibitors can improve ORR and PFS in patients with advanced or metastatic lung cancer, but the incidence of adverse reactions is high and generally tolerable. The survival of SCLC is shorter than that of NSCLC,^2^ and treatment is limited. Dual immunosuppressants also have therapeutic effects in SCLC. Despite some limitations, our research indicates that the current PD-1/PD-L1 plus CTLA-4 inhibitors may be a promising treatment strategy for patients with advanced lung cancer, but attention should be paid to the occurrence of adverse reactions.

## Data Availability

No data are available.

## Footnotes

### Contributors

XS and HX: conception and design of the study, acquisition of data, analysis and interpretation of data, drafting the article, final approval; JL and ZR: acquisition of data, analysis and interpretation of data, drafting the article, final approval; SH: interpretation of data, revising the article, final approval; BX: interpretation of data, revising the article, critical revision,final approval.

### Patient consent for publication

Not required.

### Provenance and peer review

Not commissioned; externally peer reviewed.

### Data availability statement

No data are available.

